# Gastric Alimetry^®^ improves patient phenotyping in gastroduodenal disorders compared to gastric emptying scintigraphy alone

**DOI:** 10.1101/2023.05.18.23290134

**Authors:** William Jiaen Wang, Daphne Foong, Stefan Calder, Gabriel Schamberg, Chris Varghese, Jan Tack, William Xu, Charlotte Daker, Daniel Carson, Stephen Waite, Thomas Hayes, Peng Du, Thomas L. Abell, Henry P. Parkman, I-Hsuan Huang, Vivian Fernandes, Christopher N. Andrews, Armen A. Gharibans, Vincent Ho, Greg O’Grady

## Abstract

**Objectives:** Gastric emptying testing (GET) assesses gastric motility, however is non-specific and insensitive for neuromuscular disorders. Gastric Alimetry® (GA) is a new medical device combining non-invasive gastric electrophysiological mapping and validated symptom profiling. This study assessed patient-specific phenotyping using GA compared to GET.

**Methods:** Patients with chronic gastroduodenal symptoms underwent simultaneous GET and GA, comprising a 30-minute baseline, ^99m^TC-labelled egg meal, and 4-hour postprandial recording. Results were referenced to normative ranges. Symptoms were profiled in the validated GA App and phenotyped using rule-based criteria based on their relationships to the meal and gastric activity: i) sensorimotor; ii) continuous; and iii) other.

**Results:** 75 patients were assessed; 77% female. Motility abnormality detection rates were: *GET* 22.7% (14 delayed, 3 rapid); *GA spectral analysis* 33.3% (14 low rhythm stability / low amplitude; 5 high amplitude; 6 abnormal frequency); *combined yield* 42.7%. In patients with normal spectral analysis, *GA symptom phenotypes* included: sensorimotor 17% (where symptoms strongly paired with gastric amplitude; median r=0.61); continuous 30%; other 53%. GA phenotypes showed superior correlations with GCSI, PAGI-SYM, and anxiety scales, whereas Rome IV Criteria did not correlate with psychometric scores (p>0.05).

Delayed emptying was not predictive of specific GA phenotypes.

**Conclusions:** GA improves patient phenotyping in chronic gastroduodenal disorders in the presence and absence of motility abnormalities with improved correlation with symptoms and psychometrics compared to gastric emptying status and Rome IV criteria. These findings have implications for the diagnostic profiling and personalized management of gastroduodenal disorders.

**Study Highlights:**

1) WHAT IS KNOWN

- Chronic gastroduodenal symptoms are common, costly and greatly impact on quality of life
- There is a poor correlation between gastric emptying testing (GET) and symptoms
- Gastric Alimetry® is a new medical device combining non-invasive gastric electrophysiological mapping and validated symptom profiling
2) WHAT IS NEW HERE

- Gastric Alimetry generates a 1.5x higher yield for motility abnormalities than GET
- With symptom profiling, Gastric Alimetry identified 2.7x more specific patient categories than GET
- Gastric Alimetry improves clinical phenotyping, with improved correlation with symptoms and psychometrics compared to GET

## Introduction

Chronic gastroduodenal symptoms are globally prevalent and cause a substantial quality of life and health economic burden (1,2). These symptoms are predominantly recognized in functional dyspepsia (FD), chronic nausea and vomiting syndromes (CNVS), and gastroparesis (3). However, defining and diagnosing these disorders is difficult due to their overlapping symptom and testing profiles (4,5). Identifying more specific patient phenotypes is required to inform targeted care (6).

Many current tests of gastric function are of uncertain clinical value (4,7). Gastric emptying (GET) is the most widely applied test and remains the diagnostic standard for gastroparesis (8). However, recent studies have shown that GET is non-specific, labile over time, and insensitive for neuromuscular pathologies (4,9). Therefore, while a finding of delayed emptying may be used to guide therapeutic options (10), a negative result is of dubious utility (6). Antroduodenal manometry offers a direct measurement of upper gut motility, but is invasive, not widely available, and is typically reserved for severe cases (7). Other techniques, such as barostat, drink tests, SmartPill^®^, and pyloric EndoFlip^®^ are evolving and/or applied in research contexts, but are yet to find a defined role in routine diagnostic pathways (7).

Gastric Alimetry^®^ (Alimetry Ltd., New Zealand) is a new test of gastric function that combines non-invasive body surface gastric mapping (BSGM) with concurrent validated symptom profiling (11,12). Gastric Alimetry enables high-resolution (HR) characterization of gastric myoelectrical activity, being optimized to maximally separate the weak gastric signals from noise, and achieving a critical advance over legacy electrogastrography (EGG) (13–16). The technique has been validated against antroduodenal manometry and invasive gastric electrical recordings (17,18), and has demonstrated capability to identify specific patient subgroups with gastric neuromuscular dysfunction, including with symptom correlations (11,19).

The purpose of this study was to perform a direct head-to-head comparison between Gastric Alimetry and scintigraphic GET, with the aim of comparing their relative diagnostic outcomes. We hypothesized that Gastric Alimetry testing, including specific symptom phenotypes arising from the test, would improve clinical phenotyping of patients with chronic gastroduodenal disorders compared to GET alone.

## Methods

### Study population

The study was conducted in Auckland, New Zealand and Western Sydney, Australia. Ethical approval was obtained (AHREC123; H13541), and all patients provided informed consent. Adult patients aged ≥18 years were recruited on referral to GET for diagnostic evaluation of chronic gastroduodenal symptoms. All patients had undergone clinical work-up by a specialist gastroenterologist, including upper gastrointestinal endoscopy to exclude alternative pathologies prior to scintigraphy referral. Exclusion criteria were pregnancy, breastfeeding, previous major gastric surgery (i.e. gastric resections, anti-reflux surgery, pyloric procedures and bariatric operations), or inability to undergo Gastric Alimetry due to adhesive allergies or damaged epigastric skin.

### Test Methodologies

Gastric Alimetry and scintigraphic GET were conducted simultaneously using compatible standardized test protocols (20,21). Tests were conducted in the morning after an overnight fast. Medications affecting GI motility were withheld for 48 hours prior, caffeine and nicotine were avoided on the day of testing, and glucose levels were controlled in diabetic subjects. The meal employed was a standard ∼255 kCal low-fat egg meal, radiolabelled with ^99m^Tc (20). This GET meal is smaller than the 482 kCal oatmeal bar and nutrient drink meal that has previously been used in clinical Gastric Alimetry studies (22). However, the use of the smaller GET meal was justified by a recent sensitivity analysis showing that caloric intakes of >250 kCal are sufficient to stimulate meal responses and reliably phenotype neuromuscular dysfunction using Gastric Alimetry (11).

Gastric Alimetry was conducted per current standardized methods (11,14). In brief, the system employs a stretchable array of 8×8 electrodes on an adhesive patch, coupled to a wearable Reader placed over the epigastrium (11,14). The abdominal skin was shaved and prepared using NuPrep (NuPrep, Weaver & Co, CO, USA) prior to array placement.

Recordings were performed over a fasting period of 30 minutes, followed by the test meal, then 4 hours of continuous recordings. Patients sat in a reclined relaxed position with limited movement, then transferred to the nuclear medicine table for imaging, with motion artifacts automatically corrected or rejected using onboard accelerometers and validated algorithms (14,15). Gastric Alimetry and GET tests were time-synchronized. Patients logged their symptoms into the validated Gastric Alimetry App throughout the test period (12). Symptoms of a continuous nature (nausea, bloating, upper gut pain, heartburn, stomach burn, and excessive fullness) were measured at minimum 15-minute intervals on 10-point Likert scales. Early satiation was measured immediately after meal ingestion on a similar scale, while discontinuous symptoms (reflux, belching, vomiting) were measured as discrete events (12).

Long-term symptoms, quality of life, and health psychology were also evaluated using validated questionnaires. These were the Rome IV criteria, Patient Assessment of Upper Gastrointestinal Symptom Severity Index (PAGI-SYM), Gastroparesis Cardinal Symptom Index (GCSI), Patient Assessment of Upper Gastrointestinal Disorders Quality of Life (PAGI-QoL), Patient Health Questionnaire-2 (PHQ-2), and State-Trait Anxiety Inventory-6 (STAI-6) (3,23–27).

### Data Analysis

The primary endpoint was the comparison of diagnostic yields between Gastric Alimetry and GET, with Gastric Alimetry yields considered on the basis of both the spectral phenotypes and symptom phenotypes arising from the test. Secondary endpoints were comparing the correlation between test diagnostic outcomes and patient-reported symptoms, quality of life and psychometrics.

Gastric emptying was profiled per standard clinical methods, with delayed gastric emptying defined as >10% retention at 4 hrs, and rapid emptying as <30% retention at 1 hr (20). Tests that were aborted due to vomiting were labelled as indeterminate.

Gastric Alimetry test interpretation was based on validated metrics and established reference intervals (11,16,22). Four spectral metrics are reported, each profiling a distinct feature of gastric function: the Gastric Alimetry Rhythm Index^TM^ (GA-RI; a measure of gastric rhythm stability), the Principal Gastric Frequency (the frequency associated with stable, persistent gastric activity as defined by GA-RI), BMI-adjusted amplitude, and the fed:fasted amplitude ratio (ff-AR). These metrics were used to classify gastric dysfunction as low rhythm stability (GA-RI <0.25), low amplitude (BMI-adjusted amplitude <22 µV), high sustained amplitude (>70 µV), high frequency (>3.35 cpm), and low frequency (<2.65 cpm) (22). A low ff-AR (<1.08) in isolation is not considered to define a motility abnormality, given that many healthy controls exhibit a high fasting baseline amplitude, which may affect the ff-AR calculation (22). An individual patient could have more than one spectral abnormality concurrently, except for low GA-RI which was considered an exclusive category as these patients are considered as having gastric neuromuscular dysfunction, often supported by a low amplitude and low ff-AR (11).

Symptom profiles from the validated Gastric Alimetry App were also categorized, based on previous literature, as: i) *continuous profile* (symptoms do not display variation with the meal or gastric activity); ii) *gastric sensorimotor profile* (i.e., symptoms increase within 30 minutes of the meal then decay as gastric activity reduces); iii) *other* (3,28,29). Categorization was performed by a rule-based profiling scheme as shown in **Supplementary Figure 1**, with examples of each profile shown in **Supplementary Figure 2**. Patients with spectral abnormalities and those with minimal symptoms (i.e., excessive fullness is ≤2 at 2.5 hours and all other symptoms are ≤2) on the day of testing did not undergo symptom profiling.

### Statistical methods

Data is presented as mean ± standard deviation (SD), or median (IQR). Statistical analyses were performed in Python v.3.9.7 (Python Software Foundation, https://www.python.org/). Student’s t test, analysis of variance, Mann-Whitney U or Kruskal-Wallis tests were used to compare continuous variables as appropriate and Chi-square test was used to compare categorical variables except when <5 were included in a group in which case Fisher’s exact test was used. Normality was assessed via visual inspection of Q-Q plots, and correlations between symptoms and amplitude were assessed using the Pearson correlation. Adjusted *p* values for multiple comparisons were made via the Benjamini and Hochberg method (30). A general linear model was used to assess the correlation between scintigraphy results and Gastric Alimetry® phenotypes to symptoms, anxiety, depression, and quality of life, with adjustment for age, sex, BMI and diabetes status. These data are reported as exponentiated beta coefficients and associated 95% confidence intervals (CI).

#### Sample size justification

Previous studies assessing the clinical value of legacy EGG techniques and gastric emptying scintigraphy have been performed with sample sizes ranging from 38-72 patients (31,32). Previous studies of BSGM have found significant group-level differences with samples of 43 patients and 43 controls (11). As such a minimum sample of 70 patients was estimated *apriori* for this first exploratory comparison of BSGM and GET.

## Results

### Patient Population

A total of 75 patients referred to GET for diagnostic evaluation of chronic gastroduodenal symptoms were recruited, of which 77% were female, of median age 43 (range 19 to 85 years), and median BMI 24.0 (range 16.6 to 42.1 kg/m^2^). Demographics and clinical characteristics are reported in **Table S1.** Simultaneous Gastric Alimetry and GET recordings were obtained successfully in all subjects. Prior to motility testing (i.e., on symptom-based assessment alone), 56 patients met Rome IV Criteria for CNVS (75%; with 52/56 also meeting FD criteria), 14 met FD criteria alone, and 5 did not meet either criteria, indicating a high chronic gastroduodenal symptom burden in the study cohort.

### Scintigraphy Results

Of the 75 patients, 72 completed scintigraphy tests, with 3 studies aborted due to vomiting and therefore being indeterminate. Results yielded a gastric emptying abnormality in 17 patients (23%), comprising 14 delayed and 3 rapid tests (**Figure 1A**). The average gastroduodenal symptom scores logged over the whole test did not statistically separate groups with normal vs abnormal (delayed or rapid) gastric emptying (p>0.05; **Supplementary Figure 3A-E)**. There were also no significant differences between these groups on chronic symptom scales or quality of life (p>0.05; **Supplementary Figure 3F-H**).

**Figure 1:**
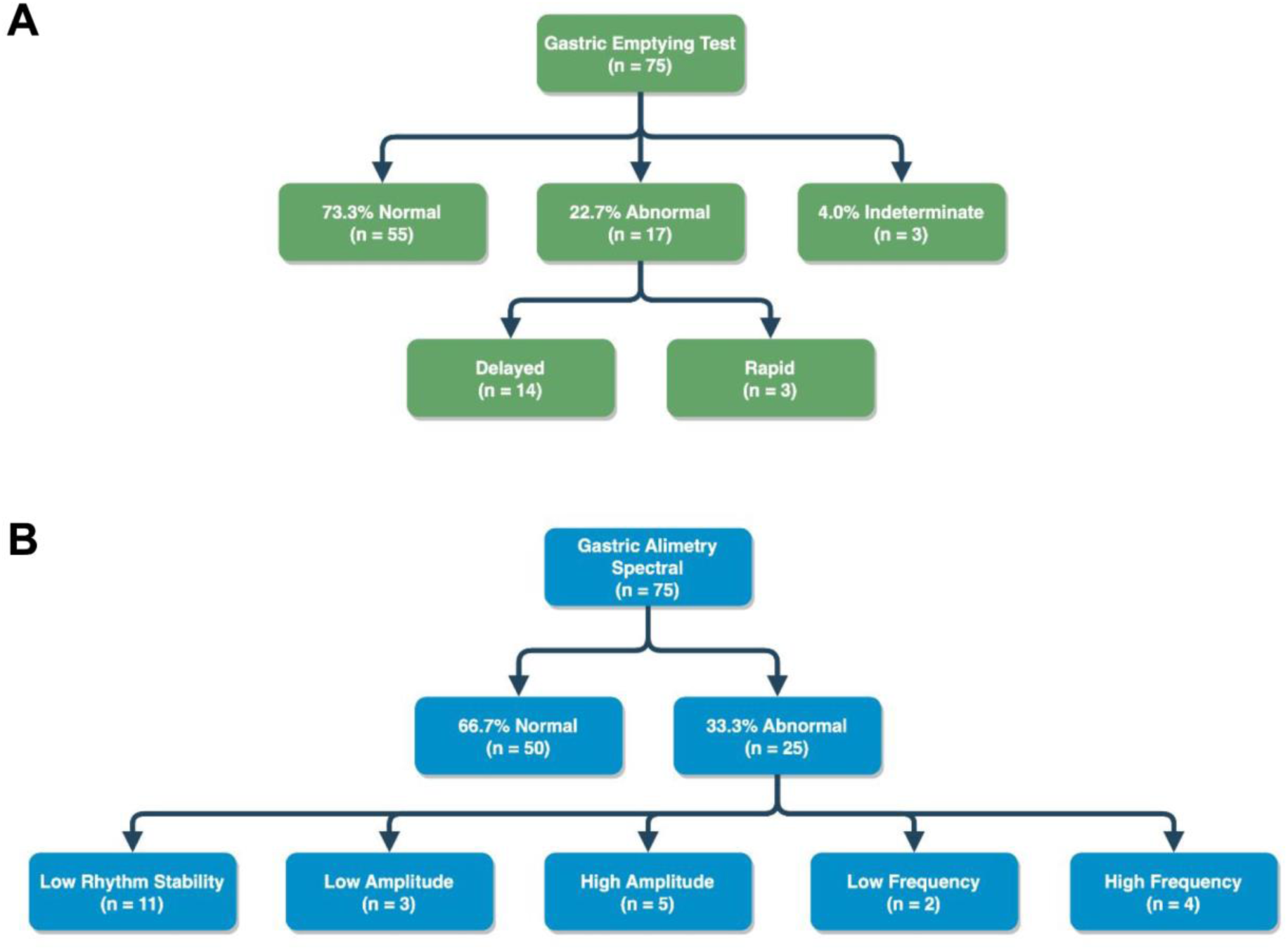
Diagnostic outcomes of GET (**A**) and Gastric Alimetry spectral analysis (**B**) per simultaneous comparison.

### Gastric Alimetry Spectral Analysis

Gastric Alimetry detected a higher rate of motility abnormalities than GET, with spectral abnormalities identified in 25 of the 75 patients (33.3%), as detailed in **Figure 1B**. These abnormalities encompassed a range of phenotypes, including low rhythm stability (low GA-RI; n=11); weak activity (low BMI-adjusted amplitude; n=3); high sustained amplitude activity (n=5); high frequency (n=4) and low frequency (n=2). Average spectrograms and amplitude curves for each phenotype are displayed in **Figure 2**. Average gastroduodenal symptoms logged over the whole test did not significantly differ between various spectral abnormalities,, except for normal vs low frequency in excessive fullness (p=0.048; **Supplementary Figure 4A-H**).

**Figure 2:**
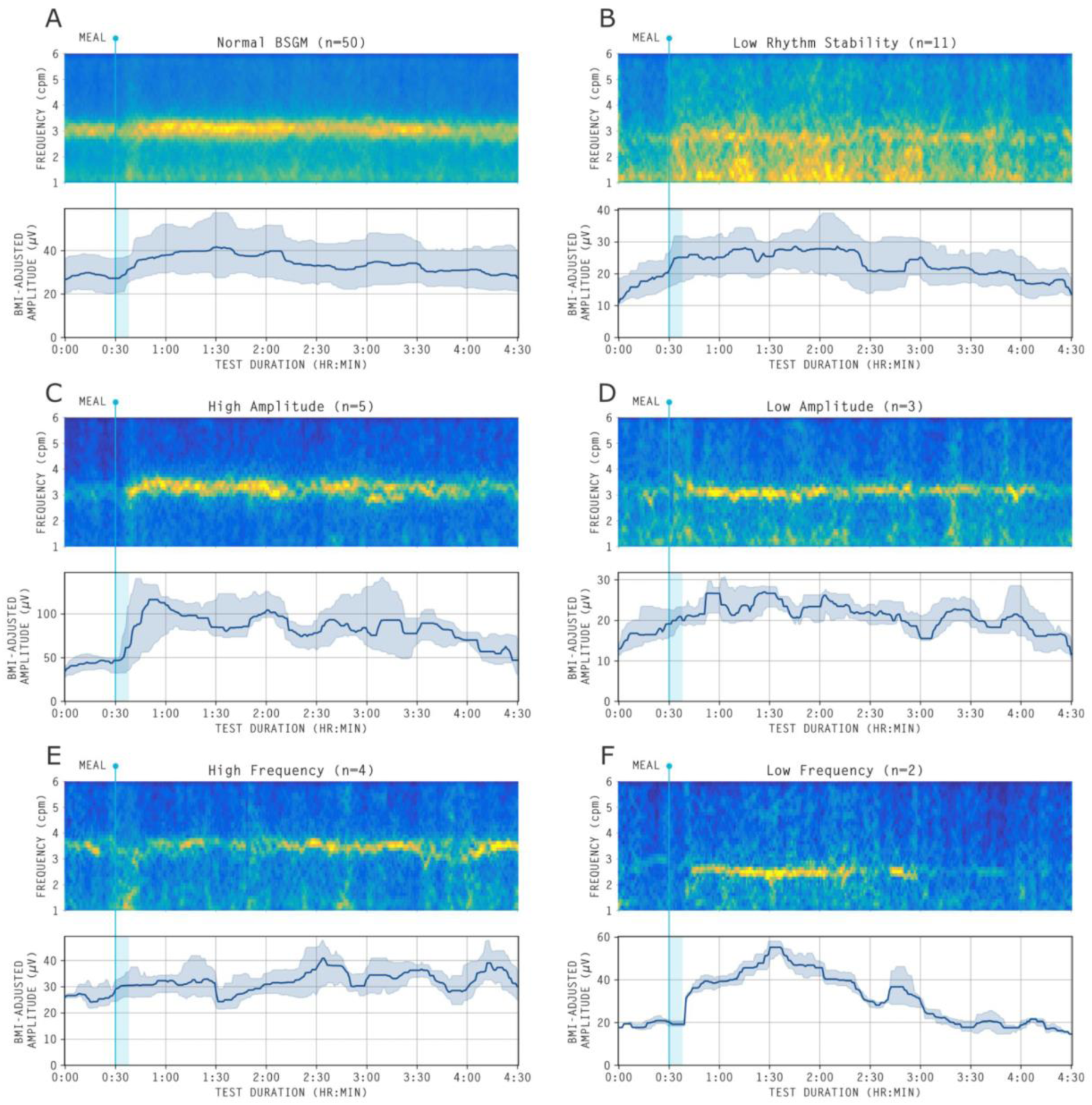
Average spectrograms for the spectral phenotypes identified by Gastric Alimetry, together with median BMI-adjusted amplitude (with 25th-75th percentiles shaded). All amplitude curves are median filtered for visual clarity. (**A**) Normal spectral analysis (n=50/75); (**B**) Low rhythm stability phenotype (low GA-RI; n=11); (**C**) High amplitude phenotype (n=5); (**D**) Low amplitude phenotype (n=3); (**E**) High frequency phenotype (n=4); **F**) Low frequency phenotype (n=2).

### Relationship between Scintigraphy and Spectral Analyses

**Figure 3A** shows the integrated outcomes of simultaneous GET and Gastric Alimetry testing. The combined diagnostic yield for any gastric motility abnormality was 42.7% (32/75), comprising 22.7% with abnormal emptying and 33.3% with Gastric Alimetry spectral abnormality, with some overlap. No associating patterns were evident between Gastric Alimetry spectral abnormalities and gastric emptying status, as seen in **Figure 3B**.

**Figure 3:**
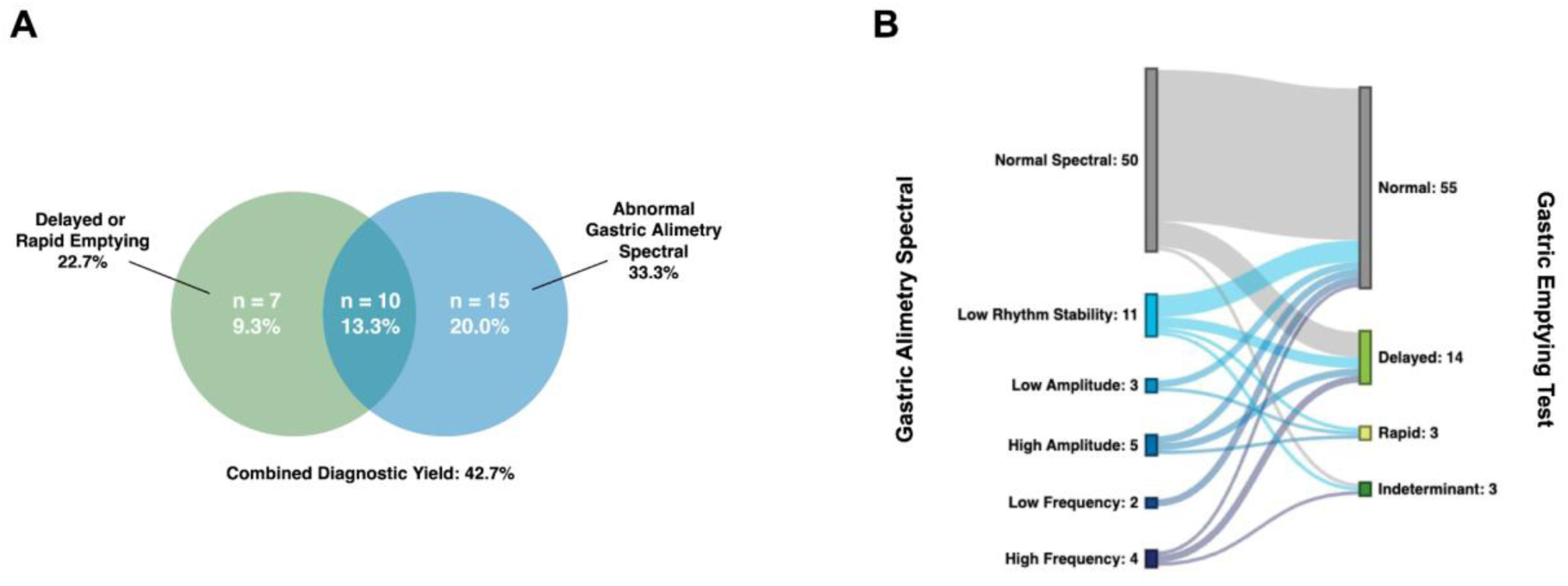
Comparison of diagnostic yields**. (A)** Comparison of a motility abnormality and their overlaps for GET and Gastric Alimetry spectral analysis; combined diagnostic yield was 42.7%. (**B)** Sankey plot showing limited concordance between gastric myoelectrical abnormalities detected by Gastric Alimetry and GET abnormalities.

### Gastric Alimetry Symptom Profiling

All 50 patients with normal Gastric Alimetry spectral analyses were considered for symptom profiling, per the scheme in **Supplementary Figure 1**. Three of these patients showed no symptoms on the day of testing and were not further profiled. Of the remaining 47 patients, symptom profiles were classified as *gastric sensorimotor* in 8 (17.0%), *continuous* in 14 (29.8%), with the remaining 25/47 classified as *other* (53.2%). Average spectrograms, amplitude curves and symptom plots for these phenotypes are displayed in **Figure 4A,B**, with the *other* group shown in **Supplementary Figure 5**. The correlation coefficient between the Gastric Alimetry log-transformed amplitude and average symptom burden profile, derived from the App during testing, was median 0.61 (IQR 0.51 to 0.65) in the gastric sensorimotor group, which was higher than that of the continuous (0.08; IQR -0.13 to 0.17; p=0.0001) and other (0.06; IQR -0.26 to 0.19; p<0.0001) groups (**Figure 4C**).

**Figure 4:**
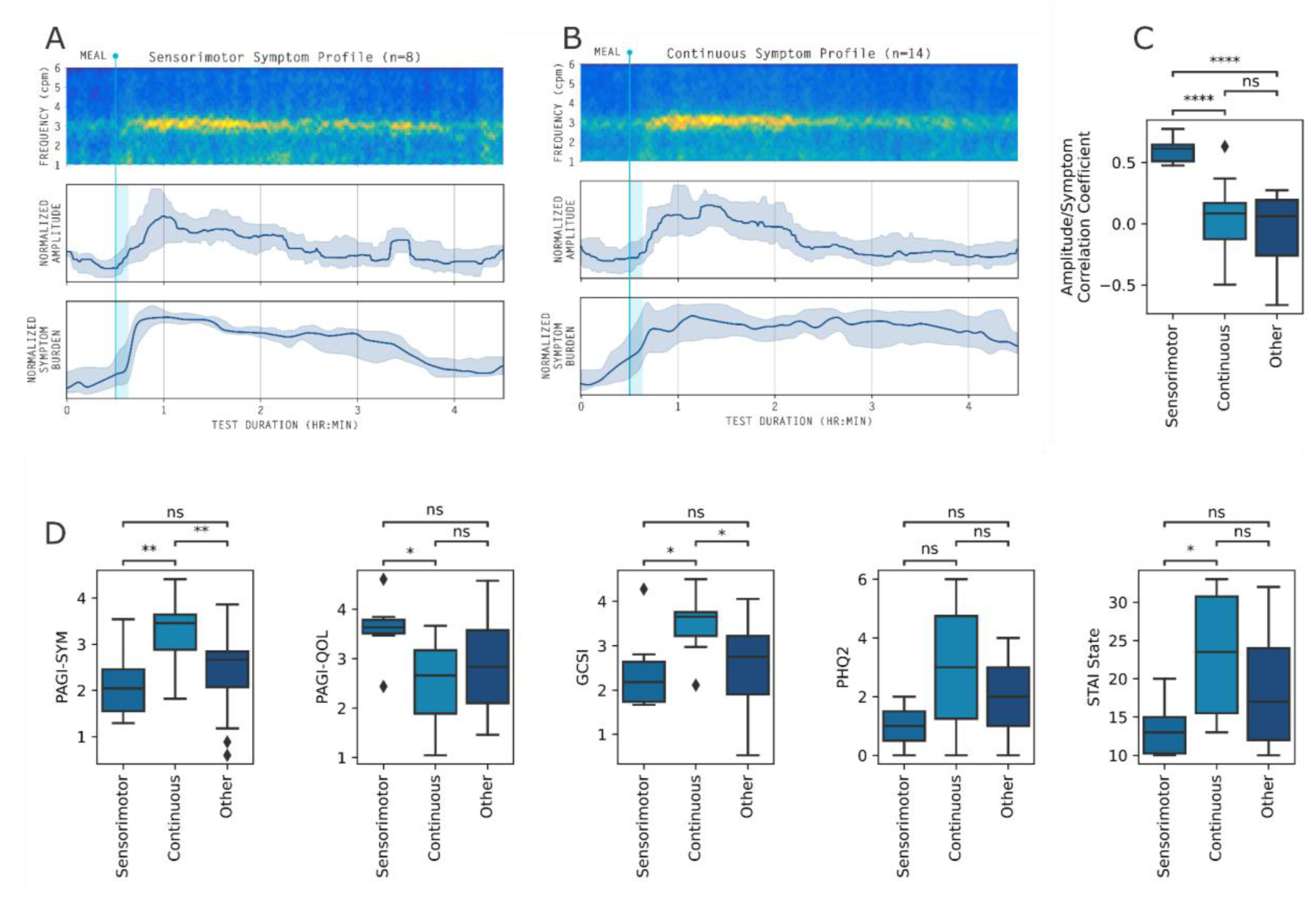
Comparison of symptom profiles. (**A)** Average spectrogram for patients with sensorimotor symptom profiles with median (IQR shaded) amplitude and symptom burden. **(B)** Average spectrogram for patients with continuous symptom profiles with median (IQR shaded) amplitude and symptom burden. **(C)** Amplitude/symptom correlation coefficients for each symptom profile. **(D)** Symptom, quality of life, and health-psychology questionnaire scores for each group. All symptom and amplitude curves normalized (mean-centered and standardized) before computing median (IQR) curves. Significance on box plots is indicated based on the result of Mann-Whitney tests with Benjamini-Hochberg correction (see Supplementary Table S2; ns: p>0.05, *: p<0.05, **: p<0.01, ***: p<0.001, ****: p<0.0001). Outliers are 1.5 x IQR from Q1 or Q3.

### Comparative correlations of Gastric Alimetry and GET with symptoms, psychometrics, and quality of life

Patients with a continuous symptom profile showed higher anxiety scores than patients with sensorimotor profiles (median STAI trait score 23.5 (IQR: 15.5 to 30.75) vs 13.0 (10.25 to 15.0); p<0.05; data unavailable for 5 patients). In addition, average symptom scores were higher in the continuous group compared to the sensorimotor group (median PAGI-SYM 3.46 (IQR 3.88 to 3.64) vs 2.04 (IQR 1.55 to 2.46); p<0.01), and quality of life was lower (median PAGI-QOL 2.66 (IQR 1.89 to 3.17). vs 3.63 (IQR 3.51 to 3.79); p=0<05). Further comparisons for questionnaire profiling of all three groups are shown in **Figure 4D** and **Supplementary Table 2**. There were no statistically significant differences in symptom scales, anxiety or depression based on gastric emptying status (**Supplementary Table 3**). These outcomes were also evaluated for Rome IV criteria (CNVS, FD including subtypes) and gastroparesis classifications for the entire cohort, as detailed in **Figure 5A**. Rome IV / gastroparesis labels including FD subtyping did not demonstrate health psychology differences (p>0.05; **Figure 5B,C)**.

**Figure 5.**
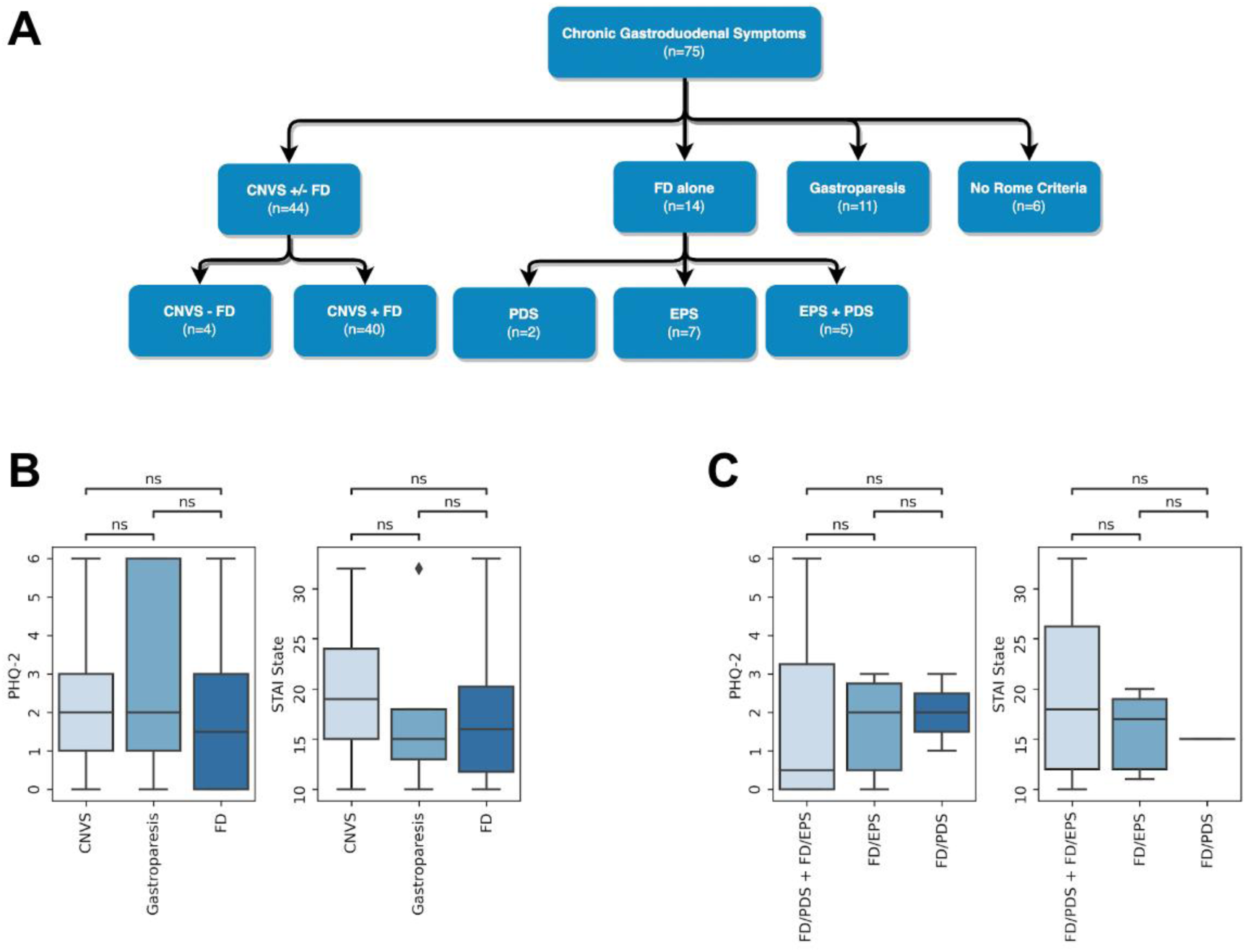
Comparison of symptoms with Rome IV criteria / gastroparesis following motility testing. (**A)** Cohort classification according to Rome IV criteria / gastroparesis (defined as predominant symptoms of nausea and delayed gastric emptying). The majority of patients had CNVS, with overlapping symptoms of with FD in 40/44 (91%). No significant differences (p>0.05) were observed in health-psychology questionnaire scores at either group-level **(B)**; or according to FD subtypes **(C)**; PDS = post-prandial distress syndrome; EPS = epigastric pain syndrome. Outliers are 1.5 x IQR from Q1 or Q3.

After adjusting for demographic confounders, symptom profiles measured by Gastric Alimetry showed better correlations with symptom questionnaires and anxiety than gastric emptying status. The continuous symptom profile was most strongly correlated with GCSI (exp(β) 4.16, 95% CI 1.99 - 8.69, p<0.001; in comparison to exp(β) 0.82, 95% CI 0.43 - 1.59, p=0.6 for delayed gastric emptying; **Table 1**). Continuous symptom profiles were most strongly correlated with anxiety (exp(β) 2.00, 95% CI 1.54 - 2.61, p<0.001), followed by the presence of Gastric Alimetry spectral abnormalities (exp(β) 1.70, 95% CI 1.36 - 2.13, p<0.001), whereas delayed and rapid emptying were not associated with anxiety (p>0.6; **Table 1**). All Gastric Alimetry phenotypes were strongly correlated with symptoms as measured by PAGI-SYM (spectral abnormality exp(β) 3.44, 95% CI 2.23 - 5.29, p<0.001, continuous profile exp(β) 7.36, 95% CI 3.72 - 14.6, p<0.001, and sensorimotor profile exp(β) 2.53, 95% CI 1.34 - 4.79, p=0.004; **Table 1**), where as delayed gastric emptying status was not correlated (p=0.4).

**Table 1:** Linear regression for assessing adjusted predictive ability of gastric emptying status and Gastric Alimetry phenotyping on GCSI, PHQ-2, State STAI, PAGI-SYM, and PAGI-QoL

|  | GCSI |  |  | PHQ-2 |  |  | State STAI |  |  | PAGI-QOL |  |  | PAGI-SYM |  |  |
| --- | --- | --- | --- | --- | --- | --- | --- | --- | --- | --- | --- | --- | --- | --- | --- |
| Characteristic | exp(Beta) | 95% CI<sup>†</sup> | p-value | exp(Beta) | 95% CI<sup>†</sup> | p-value | exp(Beta) | 95% CI<sup>†</sup> | p-value | exp(Beta) | 95% CI<sup>†</sup> | p-value | exp(Beta) | 95% CI<sup>†</sup> | p-value |
| **Age** | 0.99 | 0.98, 1.00 | 0.2 | 0.98 | 0.96, 1.01 | 0.2 | 1 | 0.99, 1.01 | >0.9 | 1.02 | 1.01, 1.03 | <0.001 | 0.99 | 0.98, 1.00 | 0.035 |
| **Sex** |  |  |  |  |  |  |  |  |  |  |  |  |  |  |  |
| F | — | — |  | — | — |  | — | — |  | — | — |  | — | — |  |
| M | 0.54 | 0.33, 0.89 | 0.016 | 0.5 | 0.22, 1.15 | 0.1 | 1.02 | 0.81, 1.28 | 0.9 | 1.46 | 0.90, 2.39 | 0.13 | 0.54 | 0.35, 0.83 | 0.006 |
| **BMI** | 1 | 0.96, 1.03 | 0.8 | 0.97 | 0.89, 1.06 | 0.5 | 1.01 | 0.99, 1.03 | 0.4 | 1.02 | 0.98, 1.06 | 0.4 | 1 | 0.96, 1.03 | 0.8 |
| **Diabetes status** |  |  |  |  |  |  |  |  |  |  |  |  |  |  |  |
| No | — | — |  | — | — |  | — | — |  | — | — |  | — | — |  |
| Type 1 | 1.7 | 1.16, 2.50 | 0.007 | 3.07 | 0.74, 12.8 | 0.12 |  |  |  | 1.81 | 0.96, 3.40 | 0.066 | 1.08 | 0.66, 1.76 | 0.8 |
| Type 2 | 2.42 | 0.72, 8.16 | 0.2 | 8.25 | 1.33, 51.3 | 0.024 | 1.28 | 0.88, 1.85 | 0.2 | 0.36 | 0.10, 1.23 | 0.1 | 2.34 | 0.79, 6.96 | 0.12 |
| **Solid scintigraphy diagnosis** |  |  |  |  |  |  |  |  |  |  |  |  |  |  |  |
| Normal | — | — |  | — | — |  | — | — |  | — | — |  | — | — |  |
| Delayed | 0.82 | 0.43, 1.59 | 0.6 | 2.06 | 0.58, 7.26 | 0.3 | 0.95 | 0.73, 1.25 | 0.7 | 0.99 | 0.58, 1.71 | >0.9 | 0.75 | 0.39, 1.43 | 0.4 |
| Indeterminant | 1.66 | 1.01, 2.73 | 0.044 | 0.30 | 0.08, 1.13 | 0.075 | 0.9 | 0.70, 1.14 | 0.4 | 1.51 | 0.79, 2.89 | 0.2 | 1.52 | 0.86, 2.68 | 0.15 |
| Rapid | 2.91 | 1.43, 5.92 | 0.003 | 5.08 | 0.35, 74.5 | 0.2 | 1.08 | 0.83, 1.39 | 0.6 | 0.25 | 0.11, 0.58 | 0.001 | 2.9 | 1.46, 5.76 | 0.002 |
| **Gastric Alimetry phenotyping** |  |  |  |  |  |  |  |  |  |  |  |  |  |  |  |
| No abnormalities detected | — | — |  | — | — |  | — | — |  | — | — |  | — | — |  |
| Abnormal spectral metrics | 1.84 | 1.05, 3.24 | 0.034 | 0.97 | 0.36, 2.09 | >0.9 | 1.7 | 1.36, 2.13 | <0.001 | 0.48 | 0.10, 2.24 | 0.4 | 3.44 | 2.24, 5.29 | <0.001 |
| Normal spectral metrics |  |  |  |  |  |  |  |  |  |  |  |  |  |  |  |
| Continuous | 4.16 | 1.99, 8.69 | <0.001 | 1.92 | 0.45, 8.24 | 0.4 | 2.00 | 1.54, 2.61 | <0.001 | 0.35 | 0.07, 1.75 | 0.2 | 7.36 | 3.72, 14.6 | <0.001 |
| Sensorimotor | 1.42 | 0.91, 3.33 | 0.4 | 0.22 | 0.06, 0.79 | 0.021 | 1.25 | 0.95, 1.65 | 0.11 | 0.93 | 0.19, 4.51 | >0.9 | 2.53 | 1.34, 4.79 | 0.004 |
| Other | 1.74 | 0.64, 3.13 | 0.095 | 1.16 | 0.38, 3.47 | 0.8 | 1.59 | 1.19, 2.13 | 0.002 | 0.37 | 0.08, 1.73 | 0.2 | 3.61 | 2.20, 5.91 | <0.001 |

<sup>1</sup> CI = Confidence Interval.

### Results Summary

A summary of results comparing scintigraphy with Gastric Alimetry spectral and symptom analysis are presented in **Figure 6**. When including all test components, scintigraphy classified 17/75 patients into disorder phenotypes (22.7%), whereas Gastric Alimetry (spectral analysis and symptom profiling) classified a higher number of patients (47/75; 62.7%; p<0.0001). Combined phenotyping of patients based on spectral analysis and symptom profiles yielded disease classifications with improved correlations to symptom scales and anxiety compared to gastric emptying status.

**Figure 6:**
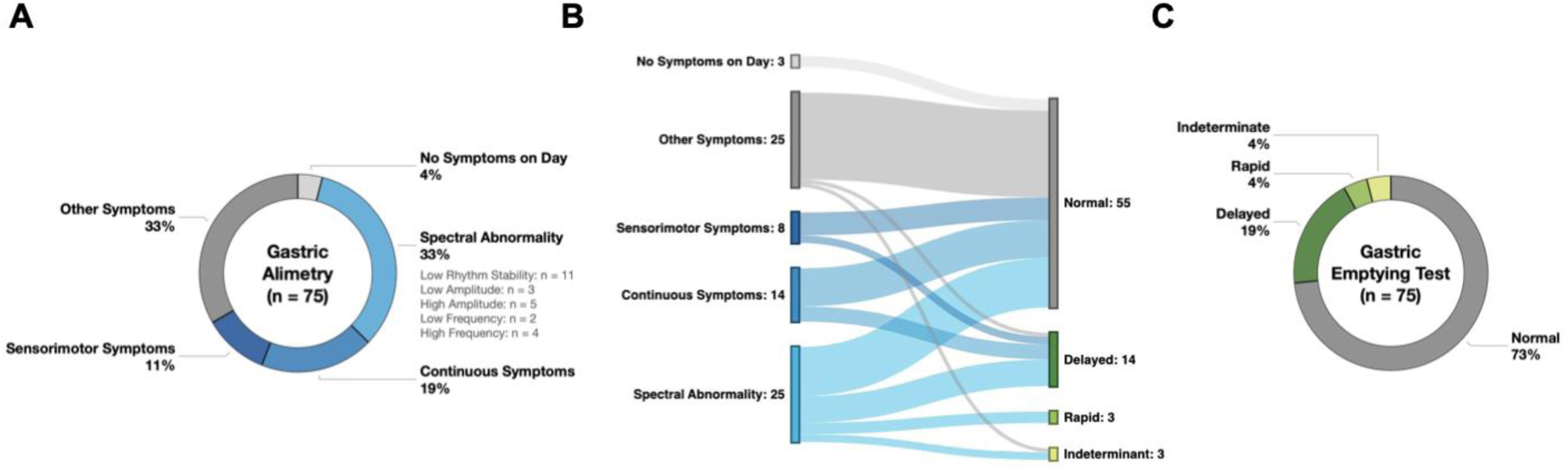
Summary of diagnostic outcomes for Gastric Alimetry vs GET. **(A)** Overall diagnostic outcomes of Gastric Alimetry spectral and symptom phenotyping. **(B)** Sankey plot showing limited concordance between Gastric Alimetry spectral or symptom phenotypes with GET abnormalities. **(C)** Overall diagnostic outcomes of GET.

## Discussion

This study performed a simultaneous comparison between Gastric Alimetry and scintigraphic GET in patients with chronic gastroduodenal symptoms. We found Gastric Alimetry spectral analysis provided a higher yield for gastric motility abnormalities *vs* GET (33.3% vs 22.7%) Gastric Alimetry also was able to further characterise patients with normal motility through additional symptom phenotypes, specifically ‘sensorimotor’ (where symptoms followed with gastric amplitude) and ‘continuous’ (which correlated strongly with anxiety and depression). Gastric Alimetry® based phenotyping correlated better with patients’ chronic symptoms and anxiety levels than gastric emptying status. Including all data, Gastric Alimetry identified 2.7x more specific patient categories than GET, with limited overlap between each diagnostic modality, offering a valuable new option in the diagnostic work up of patients with chronic gastroduodenal symptoms.

Gastric electrophysiological mapping and gastric emptying likely evaluate different aspects of gastric function, with Gastric Alimetry being sensitive to neuromuscular abnormalities, whereas GET is not (4,11). This is expected, because gastric transit is a higher-order physiological function, integrative of several other functions including autonomic activity, accommodation, peristalsis, pyloric tone, and duodenal resistance (6,33). By contrast, the electrophysiological mapping metrics assess gastric neuromuscular activity, encompassing slow waves derived from interstitial cells of Cajal (ICC), associated smooth muscle contractions, and their indirect neuronal influences (11,34). Gastroduodenal symptoms arise from diverse underlying mechanisms, and the diagnostic yield for each test was therefore within the expected range (11). It was also notable that specific symptoms alone could not predict patients with or without abnormal motility in this cohort, highlighting that physiological profiling remains essential for unravelling the neuromotor dysfunction in these disorders. Importantly, GET status could not stratify severity of symptoms, highlighting the need for better diagnostic tools to characterise the factors contributing to patient symptoms. It is also acknowledged that additional mechanisms, for example duodenal immune activation, are not profiled by either of these tests (35).

GET is widely used to inform prokinetic and pyloric therapies (36,37), however, its clinical utility as a diagnostic determinant is currently controversial. A prominent study recently showed that delayed emptying status is highly labile over time, with ∼40% of patients changing status at 48 weeks follow-up, such that FD and gastroparesis were effectively interchangeable disorders (4). In addition, delayed emptying was insensitive for neuromuscular pathologies when referenced to full-thickness biopsies, leading the authors to conclude that “gastric emptying measurements do not capture the pathophysiology adequately” (4). The development of gastric electrical mapping was motivated to address this need, by providing a direct physiological measure of gastric neuromuscular function (11,14). This has been supported by several studies showing direct associations between gastric electrophysiological abnormalities and neuromuscular / ICC pathologies, including in patients with and without delayed emptying (38–40). A separate Gastric Alimetry study recently highlighted this capability, by identifying a specific nausea and vomiting patient subgroup with gastric dysfunction, as distinct from patients with normal gastric activity but a substantially higher rate of psychological comorbidities (11).

In addition to non-invasive assessment of gastric motility, patient phenotyping is extended in Gastric Alimetry through validated symptom profiling, providing a valuable secondary layer of diagnostic data (12). This was formalized in the current study by introducing objective symptom phenotyping criteria, which provided insights into symptom origins. Symptoms in the sensorimotor phenotype were meal-responsive and showed a strong temporal correlation with gastric amplitude, likely reflecting gastric hypersensitivity or accommodation disorders (28,29). By contrast, continuous symptoms were uncorrelated with gastric amplitude but strongly associated with psychological comorbidities, indicating a gut-brain axis relationship (11,41). Furthermore, we compared these profiles to Rome IV classifications, which by contrast were not able to detect these health psychology differences. In future, the group currently labelled ‘other’ could also be further evaluated for overlaps, and to assess for additional phenotypes (21).

A large proportion of patients had conflicting results between their GET and Gastric Alimetry. In particular, almost half of the patients with delayed emptying had a normal spectral analysis (**Figure 6**). This is significant since delayed emptying is used to define gastroparesis (8). In these patients, who lacked objective evidence for a neuromuscular abnormality on BSGM, an alternative label may be more appropriate, such that they could avoid a diagnosis of gastroparesis and the serious implications this has for prognosis, therapeutic decisions, morbidity and quality of life (8,42). Similarly, about half of the patients with abnormal BSGM spectral analysis had a normal GET result. These patients are typically diagnosed with FD or CNVS but may in fact have an underlying gastric neuromuscular disorder that would otherwise go undiagnosed (4,38,39). Comparison to tissue biopsies in future would provide further clarity on these relationships, as has been achieved separately for each modality in the past (4,38–40). However, given the specificity of the BSGM for gastric myoelectrical function, and improved correlation with symptoms and psychometrics, which GET does not address, we propose that BSGM be considered the new physiological paradigm for clinically evaluating gastric neuromuscular disorders once mechanical obstruction has been ruled out with gastroscopy (11,38–40).

Several limitations of this study are noted. Although the protocols of the two tests were compatible, some compromises impacted optimal performance of the Gastric Alimetry test. Patients were required to periodically mobilize between the scintigraphy table and waiting areas, introducing motion artifacts. These were able to be corrected or rejected using validated algorithms (15,17), mitigating their significance over a 4-hr test duration. The standard scintigraphy egg meal was also of a smaller size than the oatmeal bar and ensure meal used previously for Gastric Alimetry reference interval development (22). However, we felt this was an appropriate compromise because a sensitivity analysis has previously shown that smaller caloric intakes are sufficient to adequately profile gastric neuromuscular function with Gastric Alimetry (11). We also included patients with BMI ≥35 in this study, while noting that data in this group should be interpreted with caution as per the Gastric Alimetry Guidelines. The main risk in high BMI patients is in over-estimating the low rhythm stability phenotype, due to declining signal-to-noise ratio, and the results were therefore reassuring in that no patients with BMI ≥35 had this abnormality.

In summary, this study shows that Gastric Alimetry improves the clinical phenotyping of patients with chronic gastroduodenal disorders in comparison to GET. Gastric Alimetry provided a higher yield for motility abnormalities while also introducing patient-specific symptom profiling, with improved clinical correlations. These results may enable enhanced diagnostic profiling and personalized management in gastroduodenal disorders.

## Data Availability

All data produced in the present work are contained in the manuscript

## Acknowledgements

We thank India Wallace, Gen Johnson and Isabella Pickering for their invaluable research assistance.

---

**Supplementary Figure 1.**
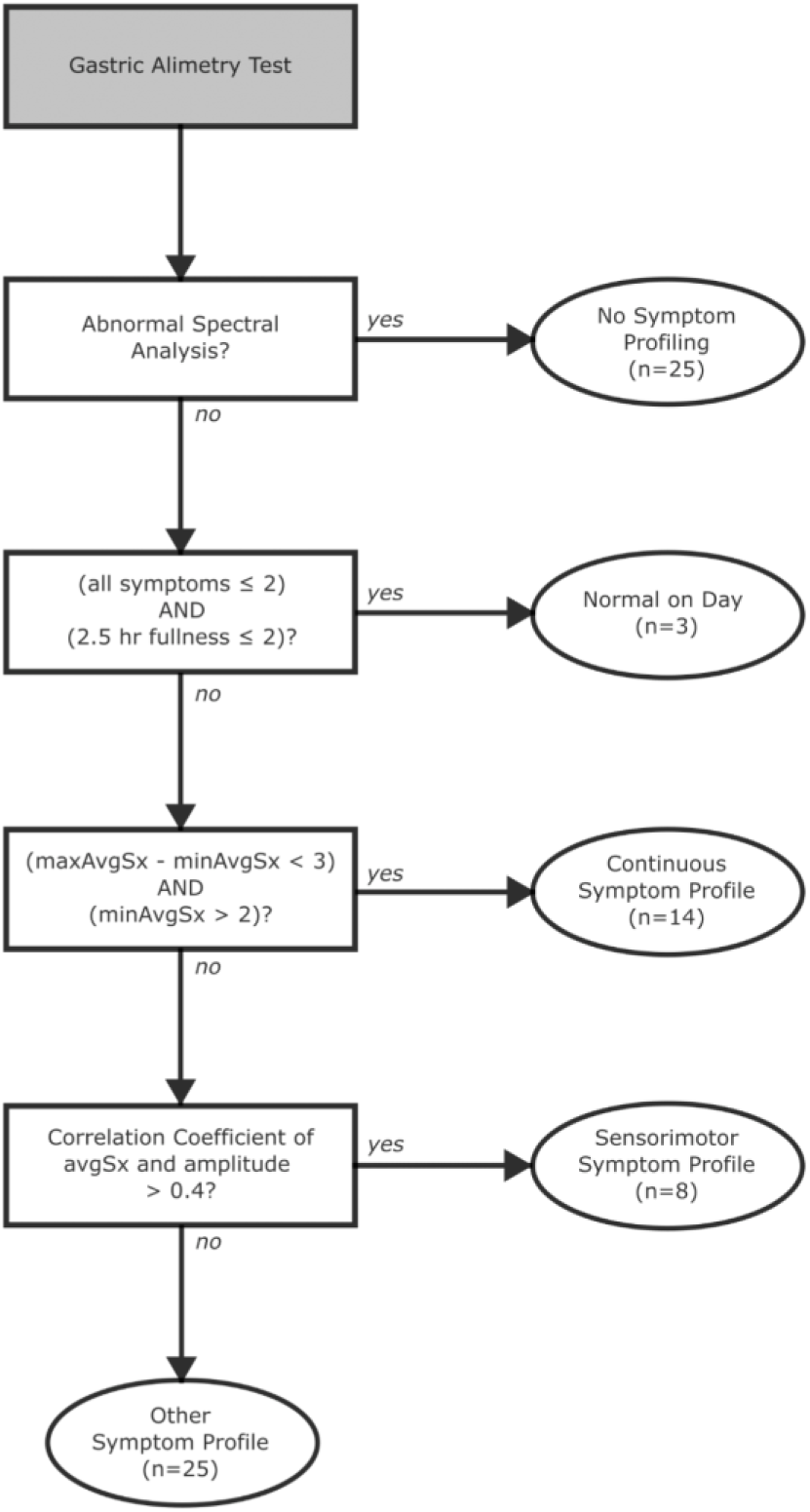
Flow chart depicting rule-based symptom profiling. Symptoms were assessed on the same day of BSGM testing and only symptoms for patients with normal BSGM spectral analysis are profiled. Patients are determined to have normal symptoms on the day if excessive fullness is ≤2 at 2.5 hours and all other symptoms are ≤2 for the whole test. Continuous symptom profiles are determined by comparing the maximum and minimum of the average symptom curve, excluding excessive fullness (maxAvgSx and minAvgSx, respectively). Sensorimotor symptom profiles are determined using the correlation coefficient of the average symptom curve (avgSx) and the amplitude curve.

**Supplementary Figure 2.**
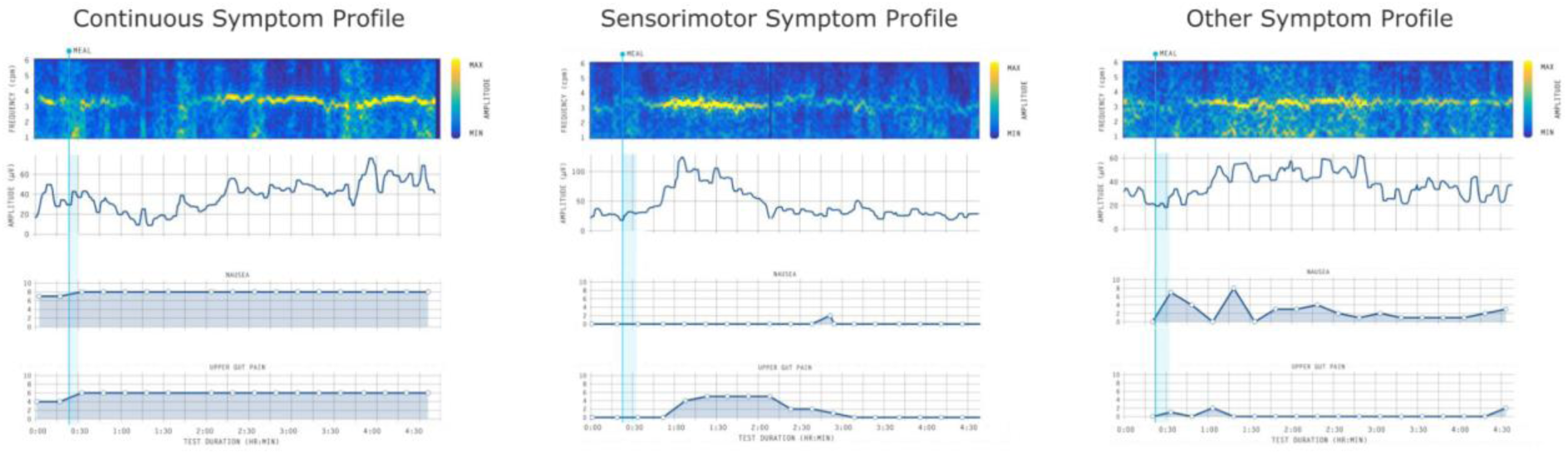
Examples of symptom profiling outcomes generated from the scheme detailed in Supplementary Figure 1.

**Supplementary Figure 3.**
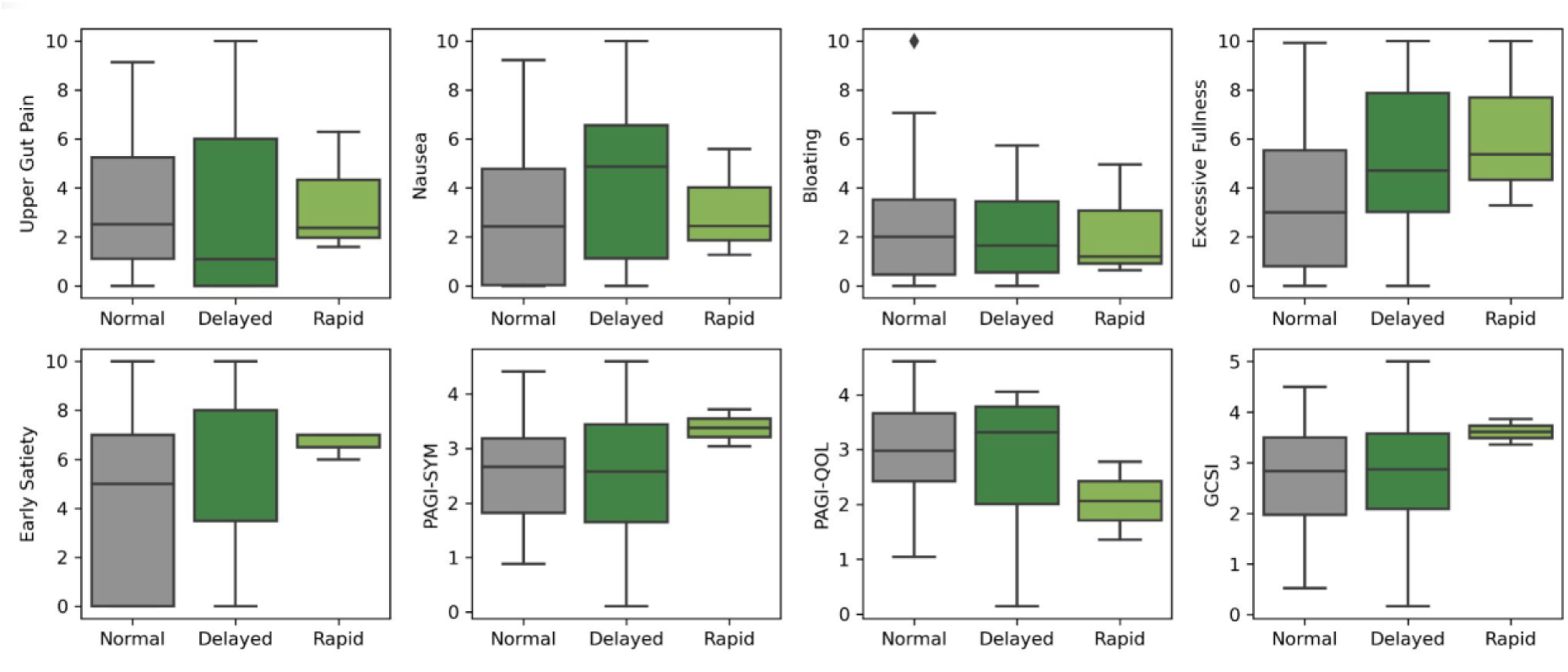
Box plots showing relationships between GET results, and symptom burdens and quality of life. Symptom burden alone did not separate subgroups. All comparisons showed p>0.05. Outliers are 1.5 x IQR from Q1 or Q3.

**Supplementary Figure 4.**
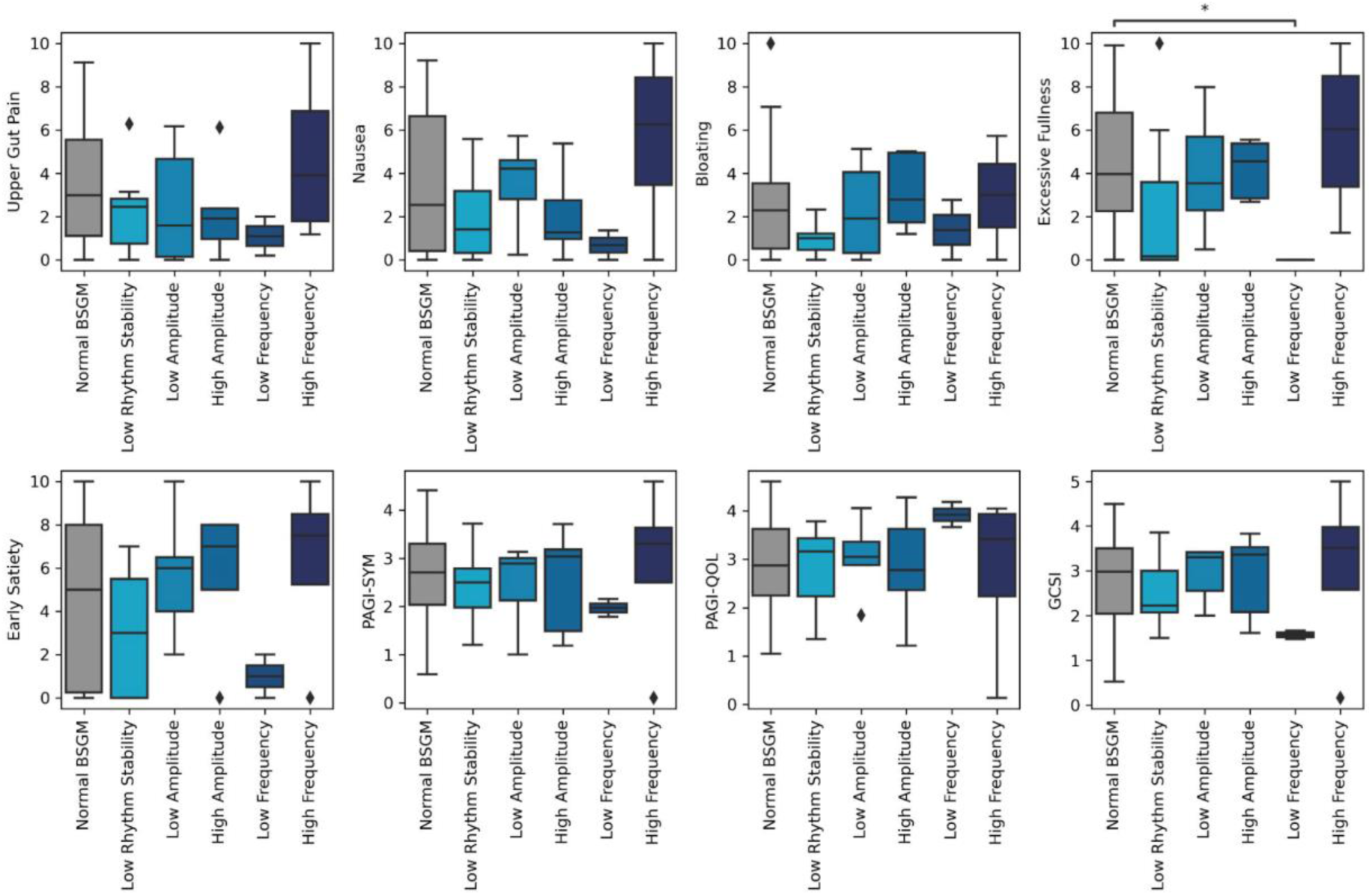
Box plots showing relationship between Gastric Alimetry spectral data,and symptom burdens and quality of life. Only normal BSGM and low frequency groups showed slight differences in excessive fullness (p=0.048); all other comparisons showed p>0.05. Outliers are 1.5 x IQR from Q1 or Q3.

**Supplementary Figure 5.**
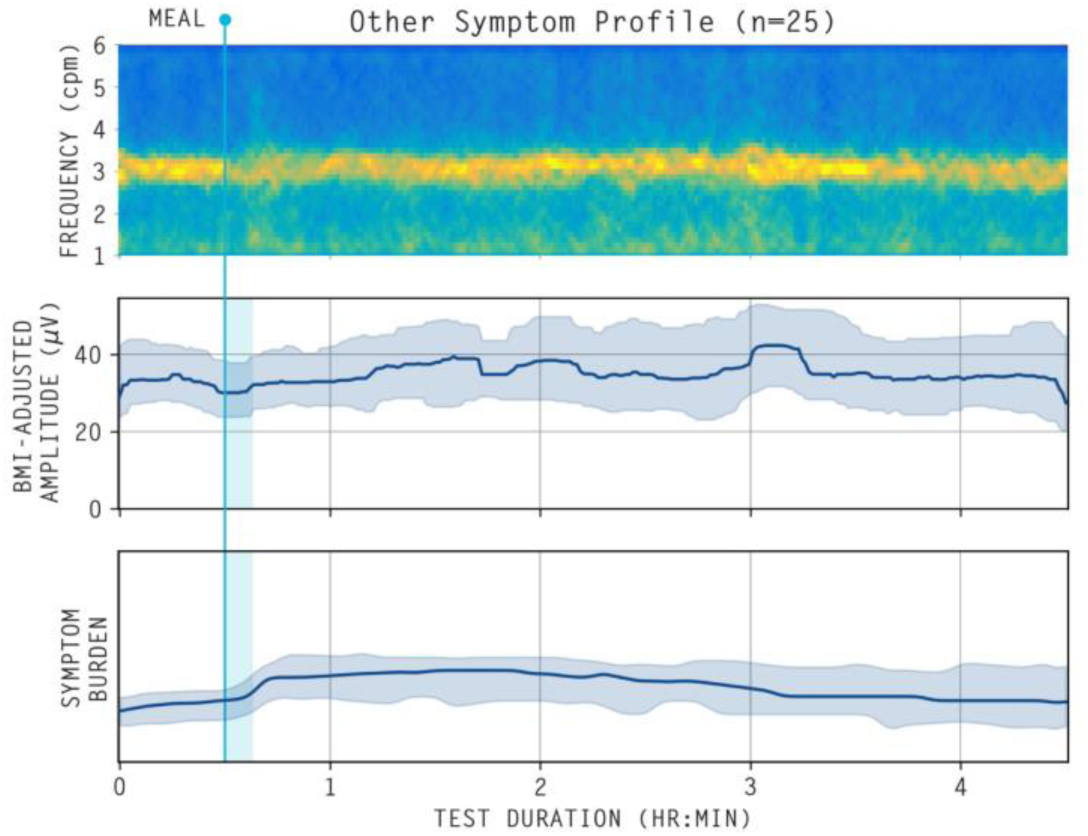
Average spectrogram, median (IQR shaded) BMI-adjusted amplitude curves, and median (IQR shaded) symptom burdens for patients with ‘Other’ symptom profiles (n=25).

**Supplementary Table 1:**
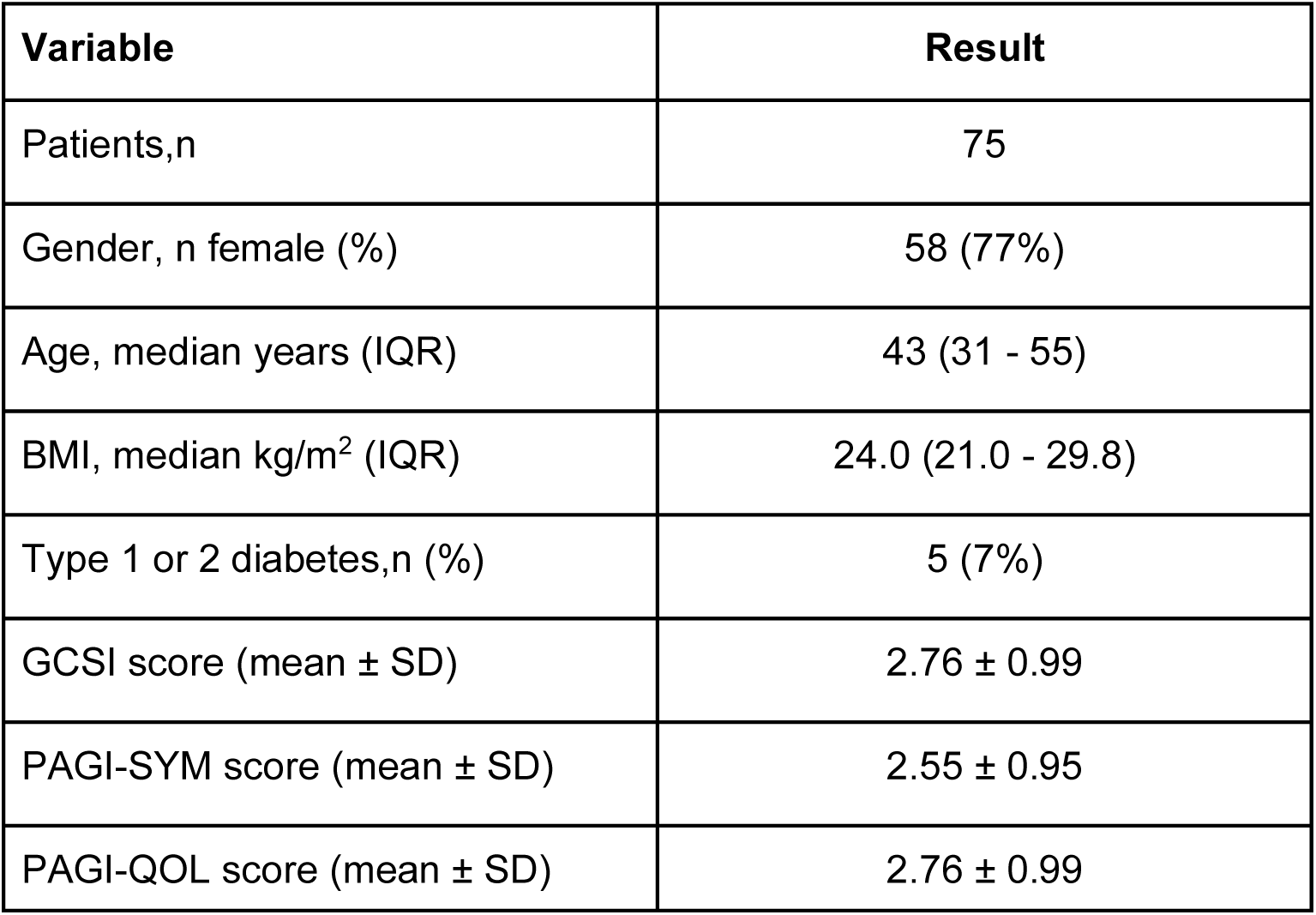
Characteristics of study cohort.

| **Variable** | **Result** |
| --- | --- |
| Patients,n | 75 |
| Gender, n female (%) | 58 (77%) |
| Age, median years (IQR) | 43 (31 - 55) |
| BMI, median kg/m<sup>2</sup> (IQR) | 24.0 (21.0 - 29.8) |
| Type 1 or 2 diabetes,n (%) | 5 (7%) |
| GCSI score (mean ± SD) | 2.76 ± 0.99 |
| PAGI-SYM score (mean ± SD) | 2.55 ± 0.95 |
| PAGI-QOL score (mean ± SD) | 2.76 ± 0.99 |

**Supplementary Table 2:** Univariate symptom and psychometric scores for patients, stratified by symptom profile.

|  | Median (IQR) |  |  |  |  | *p value* |  |  |  |  |
| --- | --- | --- | --- | --- | --- | --- | --- | --- | --- | --- |
|  | No abnormalities detected | BSGM Abnormal spectrogram | Normal BSGM; Continuous symptom profile | Normal BSGM; Sensorimotor symptom profile | Normal BSGM; Other symptom profile | Abnormal spectrogram / Continuous | Abnormal spectrogram/ Sensorimotor | Continuous / Sensorimotor | Sensorimotor / Other | Continuous / Other |
| PAGI-SYM | 1.44 (1.18 - 2.22) | 2.69 (1.80 to 3.16) | 3.46 (2.88 to 3.64) | 2.04 (1.55 to 2.46) | 2.67 (2.07 to 2.84) | **0.046** | 0.422 | **0.011** | 0.403 | **0.011** |
| PAGI-QOL | 2.86 (2.84 - 3.49) | 3.16 (2.41 to 3.72) | 2.66 (1.89 to 3.17) | 3.63 (3.51 to 3.79) | 2.84 (2.10 to 3.58) | 0.293 | 0.394 | **0.012** | 0.175 | 0.6 |
| GCSI* | 2.06 (1.94 - 2.44) | 2.83 (1.99 to 3.42) | 3.65 (3.22 to 3.76) | 2.18 (1.74 to 2.64) | 2.75 (1.90 to 3.22) | **0.026** | 0.698 | **0.026** | 0.805 | **0.016** |
| PHQ2* | 1.00 (0.50 - 2.00) | 2.00 (1.00 to 2.00) | 3.00 (1.25 to 4.75) | 1.00 (0.50 to 1.50) | 2.00 (1.00 to 3.00) | 0.241 | 0.302 | 0.079 | 0.302 | 0.241 |
| STAI State* | 11.00 (10.50 - 16.00) | 18.00 (16.00 to 21.00) | 23.50 (15.50 to 30.75) | 13.00 (10.25 to 15.00) | 17.00 (12.00 to 24.00) | 0.163 | 0.163 | **0.042** | 0.163 | 0.163 |

*p values have been adjusted for multiple comparisons using Benjamini and Hochberg correction. \*Pairwise post-hoc t tests were performed.*
*All other comparisons are made using the pairwise Wilcoxon rank sum exact test.*

**Supplementary Table 3:** Univariate symptom and psychometric scores for patients, stratified by gastric emptying status.

|  | Median (IQR) |  |  |  | *p value* |  |  |
| --- | --- | --- | --- | --- | --- | --- | --- |
|  | Normal | Delayed | Rapid | Indeterminant | Normal / Delayed | Normal / Rapid | Delayed / Rapid |
| PAGI-SYM | 2.67 (1.87 to 3.12) | 2.58 (1.47 to 3.58) | 3.38 (3.21 to 3.55) | 2.89 (2.89 to 3.10) | 0.71 | 0.6 | 0.6 |
| PAGI-QOL | 2.97 (2.38 to 3.66) | 3.23 (2.10 to 3.73) | 2.06 (1.71 to 2.42) | 2.94 (2.91 to 3.36) | 0.98 | 0.6 | 0.87 |
| GCSI* | 2.82 (1.98 to 3.42) | 2.88 (2.06 to 3.79) | 3.61 (3.49 to 3.74) | 3.31 (3.24 to 3.35) | 0.72 | 0.61 | 0.61 |
| PHQ2* | 2.00 (1.00 to 3.00) | 2.00 (1.00 to 6.00) | 3.00 (2.00 to 4.00) | 1.00 (0.50 to 1.50) | 0.32 | 0.49 | 0.91 |
| STAI State* | 17.50 (12.75 to 23.25) | 16.50 (12.50 to 24.50) | 21.00 (21.00 to 21.00) | 15.00 (14.50 to 16.50) | 0.8 | 0.8 | 0.8 |

*p values have been adjusted for multiple comparisons using Benjamini and Hochberg correction. \*Pairwise post-hoc t tests were performed. All*
*other comparisons are made using the pairwise Wilcoxon rank sum exact test.*

## References

1. Sperber AD, Bangdiwala SI, Drossman DA, et al. Worldwide Prevalence and Burden of Functional Gastrointestinal Disorders, Results of Rome Foundation Global Study. Gastroenterology 2021;160:99–114.e3.

2. Black CJ, Drossman DA, Talley NJ, et al. Functional gastrointestinal disorders: advances in understanding and management. Lancet 2020;396:1664–1674.

3. Stanghellini V, Chan FKL, Hasler WL, et al. Gastroduodenal Disorders. Gastroenterology 2016;150:1380–1392.

4. Pasricha PJ, Grover M, Yates KP, et al. Functional Dyspepsia and Gastroparesis in Tertiary Care are Interchangeable Syndromes With Common Clinical and Pathologic Features. Gastroenterology 2021;160:2006–2017.

5. Pasricha PJ, Colvin R, Yates K, et al. Characteristics of patients with chronic unexplained nausea and vomiting and normal gastric emptying. Clin. Gastroenterol. Hepatol. 2011;9:567–76.e1–4.

6. O’Grady G, Carbone F, Tack J. Gastric sensorimotor function and its clinical measurement [Internet]. Neurogastroenterol. Motil. 2022;Available from: http://dx.doi.org/to follow

7. Lacy BE, Crowell MD, Cangemi DJ, et al. Diagnostic Evaluation of Gastric Motor and Sensory Disorders. Am. J. Gastroenterol. 2021;116:2345–2356.

8. Camilleri M, Kuo B, Nguyen L, et al. ACG Clinical Guideline: Gastroparesis [Internet]. Official journal of the American College of Gastroenterology | ACG 2022;117Available from: https://journals.lww.com/ajg/Fulltext/2022/08000/ACG_Clinical_Guideline Gastropare sis.15.aspx

9. Vijayvargiya P, Jameie-Oskooei S, Camilleri M, et al. Association between delayed gastric emptying and upper gastrointestinal symptoms: a systematic review and meta-analysis. Gut 2019;68:804–813.

10. Tack J, Schol J, Horowitz M. Gastroparesis: A Dead-end Street After All? Gastroenterology 2021;160:1931–1933.

11. Gharibans AA, Calder S, Varghese C, et al. Gastric dysfunction in patients with chronic nausea and vomiting syndromes defined by a noninvasive gastric mapping device. Sci. Transl. Med. 2022;14:eabq3544.

12. Sebaratnam G, Karulkar N, Calder S, et al. Standardized system and App for continuous patient symptom logging in gastroduodenal disorders: Design, implementation, and validation. Neurogastroenterology & Motility 2022;34:e14331.[cited 2022 Sep 26]

13. Carson DA, O’Grady G, Du P, et al. Body surface mapping of the stomach: New directions for clinically evaluating gastric electrical activity. Neurogastroenterol. Motil. 2021;33:e14048.

14. Gharibans AA, Hayes TCL, Carson DA, et al. A novel scalable electrode array and system for non-invasively assessing gastric function using flexible electronics. Neurogastroenterol. Motil. 2022;e14418.

15. Calder S, Schamberg G, Varghese C, et al. An automated artifact detection and rejection system for body surface gastric mapping. Neurogastroenterol. Motil. 2022;34:e14421.

16. Schamberg G, Varghese C, Calder S, et al. Revised spectral metrics for body surface measurements of gastric electrophysiology [Internet]. bioRxiv 2022;Available from: https://www.medrxiv.org/content/10.1101/2022.07.05.22277284.abstract

17. Gharibans AA, Smarr BL, Kunkel DC, et al. Artifact Rejection Methodology Enables Continuous, Noninvasive Measurement of Gastric Myoelectric Activity in Ambulatory Subjects. Sci. Rep. 2018;8:5019.

18. Calder S, Cheng LK, Andrews CN, et al. Validation of noninvasive body-surface gastric mapping for detecting gastric slow-wave spatiotemporal features by simultaneous serosal mapping in porcine. Am. J. Physiol. Gastrointest. Liver Physiol. 2022;323:G295–G305.

19. Gharibans AA, Coleman TP, Mousa H, et al. Spatial Patterns From High-Resolution Electrogastrography Correlate With Severity of Symptoms in Patients With Functional Dyspepsia and Gastroparesis. Clin. Gastroenterol. Hepatol. 2019;17:2668– 2677.

20. Abell TL, Camilleri M, Donohoe K, et al. Consensus Recommendations for Gastric Emptying Scintigraphy: A Joint Report of the American Neurogastroenterology and Motility Society and the Society of Nuclear Medicine. J. Nucl. Med. Technol. 2008;36:44.

21. O’Grady G, Varghese C, Schamberg G, et al. Principles and Clinical Methodology of Body Surface Gastric Mapping: Technical Review [Internet]. Neurogastroenterology and Motility Available from: http://dx.doi.org/DOI to follow

22. Varghese C, Schamberg G, Calder S, et al. Normative values for body surface gastric mapping evaluations of gastric motility using Gastric Alimetry: spectral analysis [Internet]. bioRxiv 2022;Available from: https://www.medrxiv.org/content/10.1101/2022.07.25.22278036.abstract

23. Revicki DA, Rentz AM, Dubois D, et al. Gastroparesis Cardinal Symptom Index (GCSI): development and validation of a patient reported assessment of severity of gastroparesis symptoms. Qual. Life Res. 2004;13:833–844.

24. Rentz AM, Kahrilas P, Stanghellini V, et al. Development and psychometric evaluation of the patient assessment of upper gastrointestinal symptom severity index (PAGI-SYM) in patients with upper gastrointestinal disorders. Qual. Life Res. 2004;13:1737–1749.

25. Loge C de la, Trudeau E, Marquis P, et al. Cross-cultural development and validation of a patient self-administered questionnaire to assess quality of life in upper gastrointestinal disorders: the PAGI-QOL. Qual. Life Res. 2004;13:1751–1762.

26. Spielberger CD, Gonzalez-Reigosa F, Martinez-Urrutia A, et al. The State-Trait Anxiety Inventory [Internet]. RIP/IJP 1971;5[cited 2022 Oct 11] Available from: http://journal.sipsych.org/index.php/IJP/article/view/620

27. Kroenke Kurt, Spitzer Robert L. The PHQ-9: A New Depression Diagnostic and Severity Measure. Psychiatr. Ann. 2002;32:509–515.

28. Bisschops R, Karamanolis G, Arts J, et al. Relationship between symptoms and ingestion of a meal in functional dyspepsia. Gut 2008;57:1495–1503.

29. Vanheel H, Vanuytsel T, Van Oudenhove L, et al. Postprandial symptoms originating from the stomach in functional dyspepsia. Neurogastroenterol. Motil. 2013;25:911–e703.

30. Benjamini Y, Hochberg Y. Controlling the False Discovery Rate: A Practical and Powerful Approach to Multiple Testing. J. R. Stat. Soc. Series B Stat. Methodol. 1995;57:289–300.

31. Parkman HP, Miller MA, Trate D, et al. Electrogastrography and gastric emptying scintigraphy are complementary for assessment of dyspepsia. J. Clin. Gastroenterol. 1997;24:214–219.

32. Kim SW, Kim TH, Choi SC, et al. Clinical correlation between scintigraphic measurement of gastric emptying and Electrogastrography in Dysmotility like functional dyspepsia. Korean J. Gastroenterol. 2001;37:240–246.

33. Farrell MB. Gastric Emptying Scintigraphy. J. Nucl. Med. Technol. 2019;47:111–119.

34. O’Grady G, Gharibans AA, Du P, et al. The gastric conduction system in health and disease: a translational review. Am. J. Physiol. Gastrointest. Liver Physiol. 2021;321:G527–G542.

35. Wauters L, Talley NJ, Walker MM, et al. Novel concepts in the pathophysiology and treatment of functional dyspepsia. Gut 2020;69:591–600.

36. Vijayvargiya P, Camilleri M, Chedid V, et al. Effects of Promotility Agents on Gastric Emptying and Symptoms: A Systematic Review and Meta-analysis. Gastroenterology 2019;156:1650–1660.

37. Martinek J, Hustak R, Mares J, et al. Endoscopic pyloromyotomy for the treatment of severe and refractory gastroparesis: a pilot, randomised, sham-controlled trial. Gut 2022;71:2170–2178.

38. O’Grady G, Angeli TR, Du P, et al. Abnormal initiation and conduction of slow-wave activity in gastroparesis, defined by high-resolution electrical mapping. Gastroenterology 2012;143:589–598.e3.

39. Angeli TR, Cheng LK, Du P, et al. Loss of Interstitial Cells of Cajal and Patterns of Gastric Dysrhythmia in Patients With Chronic Unexplained Nausea and Vomiting. Gastroenterology 2015;149:56–66.e5.

40. Lin Z, Sarosiek I, Forster J, et al. Association of the status of interstitial cells of Cajal and electrogastrogram parameters, gastric emptying and symptoms in patients with gastroparesis. Neurogastroenterol. Motil. 2010;22:56–61, e10.

41. Xu W, Gharibans AA, Calder S, et al. Defining and phenotyping gastric abnormalities in long-term type 1 diabetes using body surface gastric mapping [Internet]. Available from: http://dx.doi.org/10.1101/2022.08.10.22278649

42. Dudekula A, O’Connell M, Bielefeldt K. Hospitalizations and testing in gastroparesis [Internet]. Journal of Gastroenterology and Hepatology 2011;26:1275– 1282.Available from: http://dx.doi.org/10.1111/j.1440-1746.2011.06735.

